# Chemical and Biological Characteristics of PM₁-Associated Aerosols and Airborne Viruses in Hospital and Campus Environments during the Post-COVID Period

**DOI:** 10.64898/2026.01.21.26344567

**Authors:** Yu-Hsiang Chen, Po Jui Chen, Kun-Ta Chou, Hsiang-Ling Ho, Kai Yu Hsu, Kung Hung Chieh, Ta-Chih Hsiao, Mey-Jy Jeng, Tse-Min Wei, Hsiao-Chi Chuang, Kai Hsien Chi

## Abstract

We utilized traditional aerosol sampling to collect PM_1_ samples, and further apply redundancy analysis (RDA) to investigate the association of environmental factors (including PM_1_ chemical composition, oxidative potential, meteorological factors and gaseous pollutants) and airborne bacterial community. Our findings revealed that *Bacteroidota* positively correlated with Sn and Mn, *Firmicutes* with local primary pollutants, and *Proteobacteria* with transportation-related pollutants. Variance partitioning analysis (VPA) showed that PM_1_ chemical composition, meteorological factors and gaseous pollutants collectively explained up to 43.7% of community variance, and synergistic effects may exist among the three factors. In contrast, oxidative potential had minimal influence.

Additionally, to investigate airborne viral presence, we employed a novel bioaerosol sampler targeting SARS-CoV-2 in hospital and campus settings. Viral loads were highest in negative pressure isolation rooms, followed by general hospital and campus areas. Also, the detection rates follow the same pattern, which is 87.5%, 58.3%, and 25.0%, respectively. Notably, detection rates near isolation wards increased during patient admissions, implying possible biocontamination despite containment measures. Peak human traffic flow emerged as a significant factor influencing viral detection. These results highlight how environmental factors shaping airborne microbial communities.

## 1. Introduction

Air pollution is one of the major challenges that modern society would confront. Among the factors that could harm human health, particulate matters (PM) or aerosols play a critical role. Physically, they could easily enter and deposit at the human respiratory tract [1–3]. Chemically, many chemical species could attach on them and cause deeper damages to human body [4–6]. Biologically, they could also carry biologic materials such as bacteria or viral particles, directly causing diseases. For example, aerosolized *Legionella pneumophila* can cause Legionnaires’ disease from air conditioning systems [7], while *Mycobacterium tuberculosis*, the pathogen responsible for tuberculosis, can also be transmitted via aerosols [8]. Similarly, viruses such as influenza virus can spread through both droplets and aerosols [9]. The notorious smallpox caused by variola virus is another example of airborne diseases [10].

During the COVID-19 pandemic, surveillance of aerosolized viruses had been confirmed feasible and have the potential for disease control [11, 12]. The surveillance could provide early warnings of viral existence, identify transmission hotspots, and contribute to a better understanding of airborne transmission pathways. Moreover, it may provide a framework for monitoring other respiratory pathogens in future outbreaks, thereby enhancing preparation and response strategies for infectious disease control.

When sampling atmospheric PM, filters (made from glass fiber or quartz) are often used as collecting materials. According to PM size, filters with an appropriate pore size are selected for sampling, typically with pore size around several micrometers. However, viral particles measure only a few tens of nanometers, which is much smaller than the pore size. As a result, it is unavailable to utilize traditional aerosol sampling technique to capture environmental viruses. To address this limitation, novel bioaerosol sampling technique specifically designed for viruses has been developed to assess environmental viral load.

This study explores the research potential of using traditional aerosol sampling to assess airborne bacterial community and novel bioaerosol sampling technique to detect environmental viruses. For bacteria, correlation between bacterial community and PM characteristics is analyzed. Several environmental factors are assessed for their ability to impact on bacterial community structure. For viruses, with consideration of the impacts of the COVID-19 pandemic, the spatial distribution of SARS-CoV-2 in hospital and campus areas is investigated. In addition, the relationship between viral load and environmental conditions is also explored.

## 2. Materials and methods

### 2.1 Sampling sites

The sampling site for traditional atmospheric suspended particle is located in the campus of National Taiwan University (NTU), beside Keelung Road (25°01’02.3’N 121°32’38.4’E), which is one of the busiest roads in Taipei City. NTU station is responsible for several science research, including air quality surveillance and air pollution analysis etc., so it was chosen as the sampling site [13, 14]. The samples were collected continuously on January 3^rd^ to 15^th^, 2024. The sampling period was 8 hours for each sample, with 6 am to 2 pm, 2 pm to 10 pm and 10 pm to 6 am of the next day being the sampling time. 35 samples were collected in total.

As for novel bioaerosol sampling, the sampling location includes hospital area and campus area. The sampling hospital is a national first-class medical center located in Taipei City, Taiwan. In this study, the sampling sites include (i) negative pressure isolation rooms of COVID-19 patients, (ii) conference room of nurse station adjacent to the negative pressure isolation rooms, and (iii) hospital lobby. The sampling period is from January to March, 2024. All samples were collected by air sampler for continuous 12 hours except one sample of negative pressure isolation room, which only sampled for 8 hours.

For campus area, two campuses were involved. One was NTU campus, the site was same as previous mentioned. The sampling period is on January 2024. Each sample was collected for 12 hours. The other sampling school is a public research university located in suburban of Taipei City, Taiwan, and is near the sampling hospital. The sampling sites include (i) convenience store inside the campus, (ii) school cafeteria, (iii) shoe removal area of swimming pool inside the campus, (iv) café inside the campus, and (v) school mail room. The sampling period is from March to May, 2024. Besides the sites mentioned above, there was a two-day international conference held in the campus during the sampling period, so we further collected two samples on the conference. The sampling duration varied from 7 to 12 hours based on human traffic conditions.

### 2.2 Sampling instruments and chemical analysis

Traditional atmospheric aerosol sampling for collecting PM_1_ and bacteria samples were using DHA-80 high volume aerosol sampler (Digitel Elektronik AG, Volketswil, Switzerland), equipped with 150 mm TissuQuartz™ quartz filters (Pall Life Sciences, Port Washington, NY, USA). The sampling flow rate was set to 500 liters per minute. For novel bioaerosol sampling collecting viral particles, AerosolSense™ Sampler (Thermo Fisher Scientific, Waltham, MA, USA) was utilized. This air sampler ran at a constant flow rate of 200 liters per minute. The cartridge contains collection substrate, which is non-hazardous, proprietary polymer material, and is plastic in nature, inert and not corrosive.

The collected samples underwent pretreatment for further chemical analysis, including water-soluble ions (WSIs), metal elements, and carbon. Ion chromatograph (IC) was performed to determine the mass concentration of selected ions using Dionex™ ICS-1000 Ion Chromatography (Thermo Fisher Scientific, Waltham, MA, USA). The analyzed WSI species includes Na^+^, NH_4_^+^, K^+^, Mg^2+^, Ca^2+^, Cl^-^, NO_3_^-^ and SO_4_^2-^. For metal elements, NexION^®^ 300X Inductively Coupled Plasma Mass Spectrometry (ICP-MS) (PerkinElmer, Waltham, MA, USA) was used to analyze the mass concentration of Al, Na, Mg, K, Ca, Sr, Ba, Ti, Mn, Zn, Mo, Sn, Sb, Pb, V, and Zr. Organic carbon and elemental carbon (OC and EC) was analyzed followed the Thermo-Optical Reflection (TOR) method established by the U.S. Environmental Protection Agency (EPA). The abovementioned chemical analyses were conducted at the Research Center for Environmental Changes of Academia Sinica, Taiwan.

Besides, we measured the oxidative potential (OP) of PM samples using dithiothreitol (DTT) assay [15, 16]. DTT assay is an acellular method to measure OP, and is particularly preferred to screening for toxicity of PMs, thus becoming the most commonly used technique in quantifying OP of PMs [17–19].

The DTT assay is based on the study of Fang, Verma [20]. In detail, the PM samples were water extracted three times by ultrasonic bath, then the extract was portioned to 1 mL centrifugal tubes. 1 mL of potassium phosphate buffer (pH=7.4) and 0.1 mL of DTT (0.5 mM) were added to the centrifugal tubes and put into 37 ℃ water bath with continuous shaking for incubation. At fixed timepoint (2, 8, 15, 22, 28 min), the centrifugal tubes were retreated in order and were added with 0.5 mL of 0.2mM of 5,5-dithiobis-(2-nitrobenzoic acid) (DTNB) (Sigma-Aldrich, Burlington, MA, USA). Reaction between DTT and DTNB generates a light-absorbing product, 2-nitro-5-thiobenzoicacid (TNB), which could be detected by ultraviolet–visible spectrophotometry (UV-Vis) at a wavelength of 412 nm. The UV-Vis we used is Evolution 200 (Thermo Fisher Scientific, Waltham, MA, USA). Absorbance intensity was recorded and linearly regressed. The rate of DTT consumption was determined by the intercept and slope of linear regression of measured absorbance versus time, showing as follows:

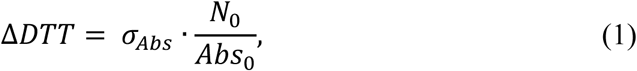

where σ_Abs_ is the slope of absorbance versus time, N_0_ is the initial moles of DTT, and Abs_0_ is the intercept of absorbance versus time. Furthermore, OP normalized by volume and mass could be calculated by formula (2):

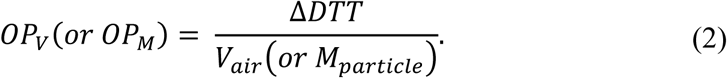

### 2.3 Bacterial identification and viral load detection

The isolation of bacterial DNA was accomplished by the use of QIAamp DNA Kits (QIAGEN, Hilden, Germany), following the manufacturer’s guidelines. The V3 and V4 region of 16S rRNA genes were sequenced and amplified for taxonomic classification. DADA2 package within R environment was applied to identify amplicon sequence variants (ASVs) and DECIPHER package was used to alignment of multiple sequences to ASVs. As a result, 386 ASVs were identified.

Viral load detection was achieved by RT-qPCR (quantitative reverse transcription polymerase chain reaction) technique since SARS-CoV-2 is RNA virus. The output value is cycle threshold (C_t_ value), with the lower C_t_ value indicating higher viral load. This work is done at Department of Pathology and Laboratory Medicine of Taipei Veterans General Hospital.

### 2.4 Data analysis

Redundancy analysis (RDA) is a commonly used method for analyzing the relationship between biological species data and environmental factors, such as chemical compound or meteorological factors [21, 22]. RDA can be seen as the combination of multivariate multiple regression and principal component analysis (PCA). In RDA, two datasets are required, namely species data and environmental variables, which serve as dependent and independent variables, respectively.

In detail, before conducting the RDA technique to the bacterial species data and selected environmental factors, all singletons were removed. In addition, Hellinger transformation was applied to bacterial species data to turn absolute abundance into relative abundance. After RDA procedure, variance inflation factor (VIF) of each environmental variable was examined, and those variables with VIF > 20 were removed from the model to prevent collinearity among independent variables. This work was done by vegan package within R environment.

## 3. Results and discussion

### 3.1 Chemical composition and temperature distribution of PM_1_ samples

Table 1 shows the mean concentration of PM_1_ chemical species, including ion, metal, carbon and OP. The PM_1_ mass concentration ranged from 3.19 to 38.02 μg/m^3^, and the temperature fluctuated from 11.7 to 22.2 ℃ (Fig. S1). No precipitation was observed during the sampling period.

Among 35 samples, PM_1_ mass concentration is a little higher in afternoon, yet no significant difference exists. On the other hand, the temperature is significantly higher in afternoon. This might cause stronger photochemical reaction and generate more secondary pollutants such as PM_1_ [23–25]. The result is shown in Fig. 1.

**Fig. 1.**
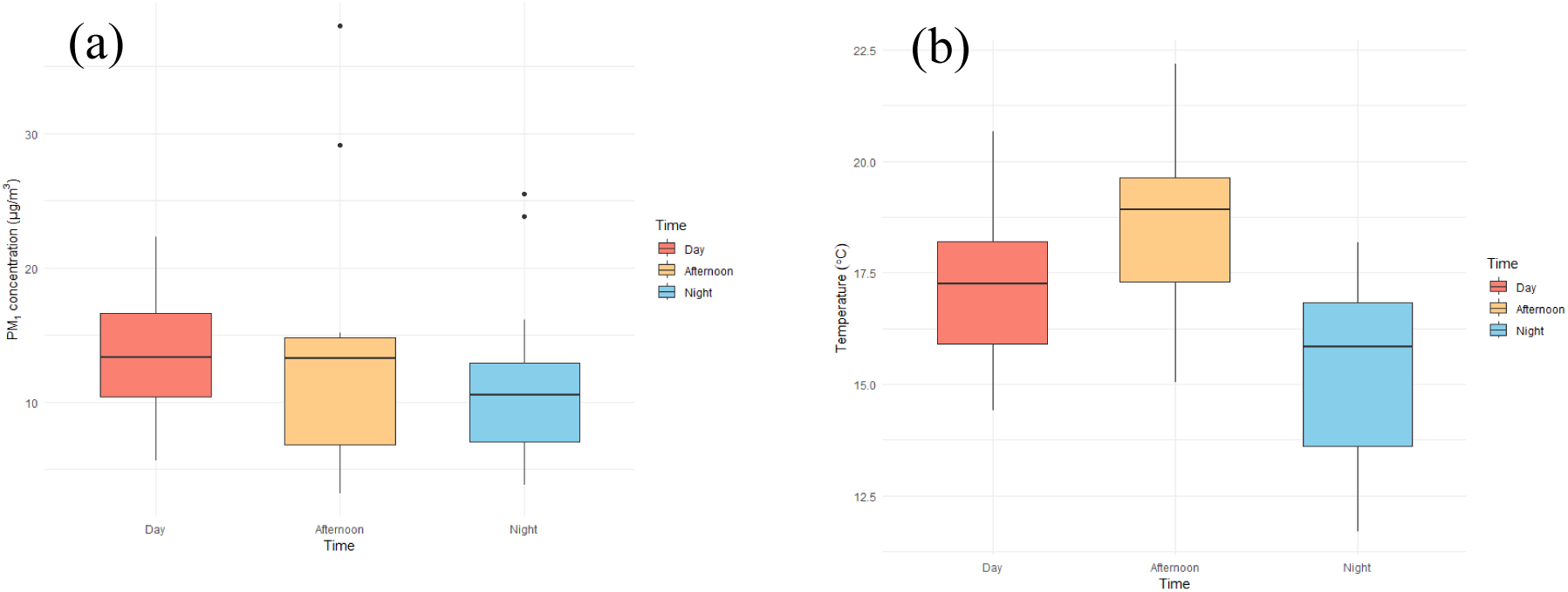
The distribution of (a) PM_1_ and (b) temperature in day, afternoon and night samples. The higher temperature in afternoon might yield higher PM_1_ concentratio

Fig. 2 shows the ion and metal species distribution of PM_1_ samples in day, afternoon, and night period. For ions, SO_4_ ^2-^ accounts for around half of total ions, while Cl^-^ and NO_3_^-^ constitute most of the remaining part. Besides, crustal elements such as Ca, Al, and K form the majority of metals in PM_1_ samples. In analysis of similarity (ANOSIM), no significant difference was observed between day, afternoon, and night samples (Fig. S2). Additionally, the temporal distribution of OC/EC and OP is shown in Fig. S3, while ANOVA also suggests no significant difference exists within a day.

**Fig. 2.**
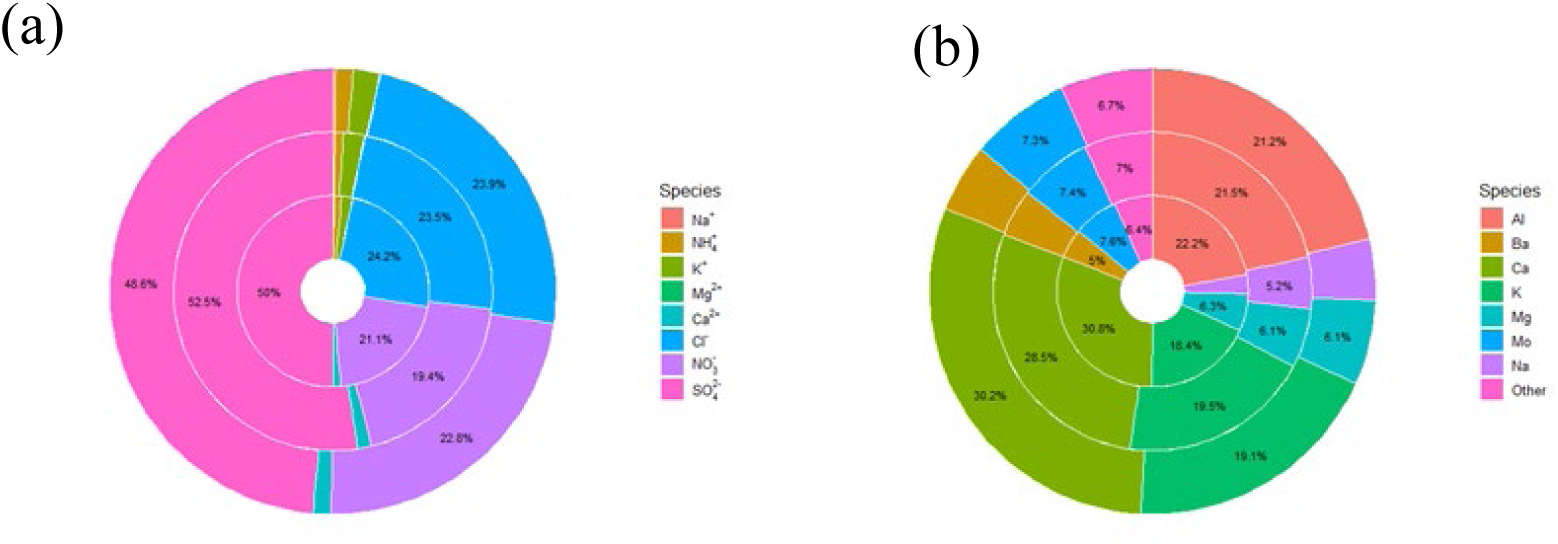
Concentric ring charts showing the distribution of (a) ionic species and (b) metal species during daytime, afternoon, and nighttime sampling periods. The inner, middle, and outer rings represent day, afternoon, and night samples, respectively

For bacterial community structure, we found that *Bacteroidota*, *Firmicutes* and *Proteobacteria* contribute to over 90% of the bacterial phyla sampled, which are common in terrestrial and aquatic environments [26, 27]. This result appears to be in accordance with other studies [27–29]. Fig. 3 shows the bacterial community in phylum levels of the samples.

**Fig. 3.**
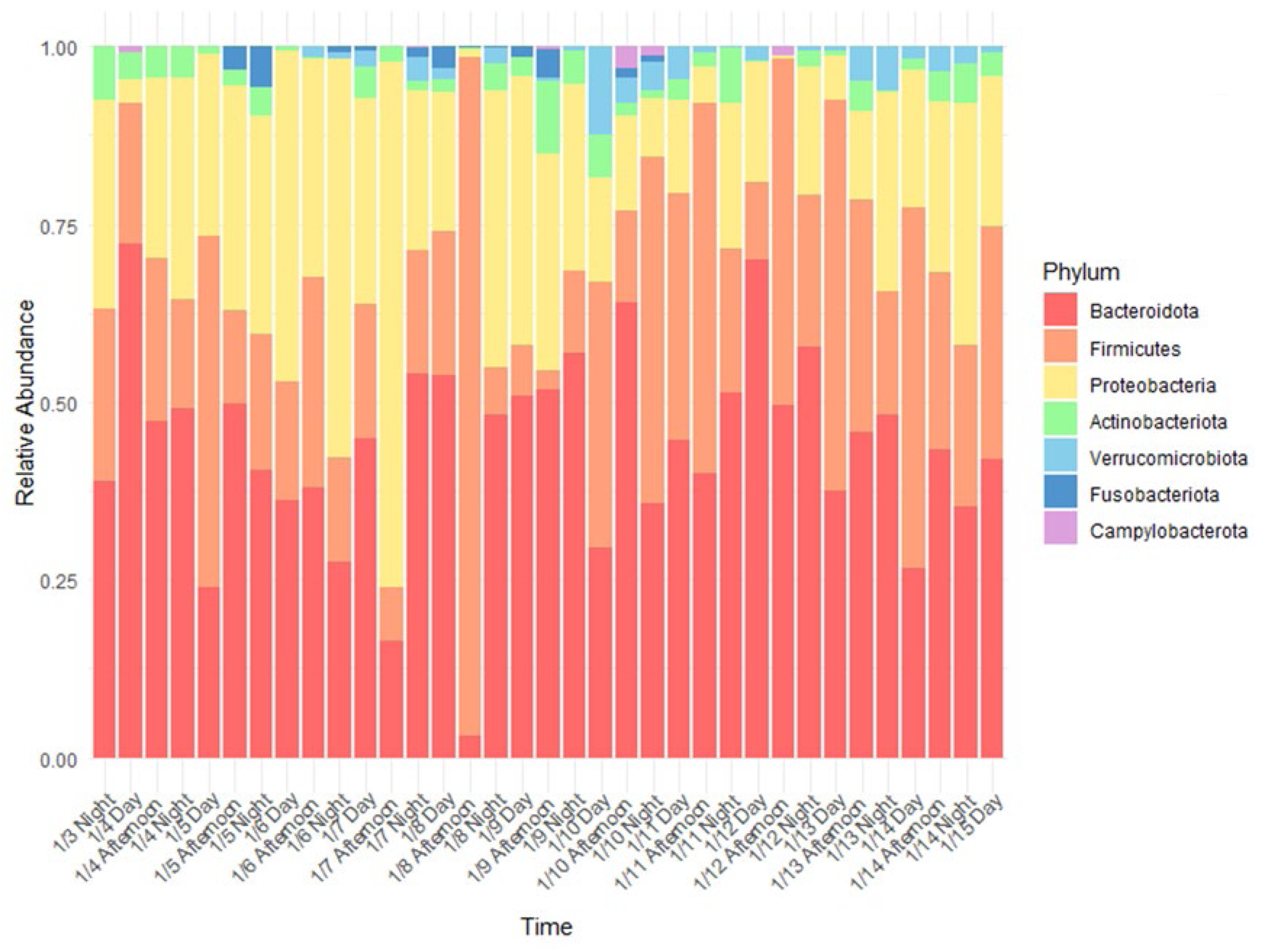
Bacterial community in phylum levels. *Bacteroidota*, *Firmicutes*, and *Protobacteria* account for over 90% of total bacteria in every sample

Compared to ions, metals and PM_1_ mass concentrations, the diurnal change of bacterial community is more obvious. This diel variation might due to fluctuation of temperature and relative humidity within a day, and in this study, temperature is a more probable factor [30]. Additionally, the diurnal change of boundary layer thickness may also reshape near-surface bacterial composition, since atmospheric turbulence is an important factor to affect microbial dispersal [31]. However, the diel change of bacterial community is not statistically significant in this study, so more research is still needed.

### 3.2 Relationship between air quality and bacterial diversity and abundance

We calculated Shannon index of each sample, which is a widely used metric to describe the diversity within a community [32]. We sought to investigate whether air quality could affect air bacterial diversity, so we also collected the air quality index (AQI) data of Guting surveillance site (25°01’14.2’N 121°31’46.4’E), which is the nearest one from our sampling site. The air quality data is provided by Taiwan Ministry of Environment (MOENV) Opendata Platform (https://data.moenv.gov.tw/dataset/detail/aqx_p_488).

We conducted linear regression analysis to assess the correlation between AQI level and Shannon index. At the phylum level, no correlation was observed (R^2^ = 0.01) (Fig. 4a). Considering the phylum-level Shannon index may be too broad-scale, we further examined the ASV-level Shannon index. Still, the analysis also revealed no correlation (R^2^ < 0.01) (Fig. 4b). These findings are inconsistent with previous studies conducted in Chile and China [33, 34].

**Fig. 4.**
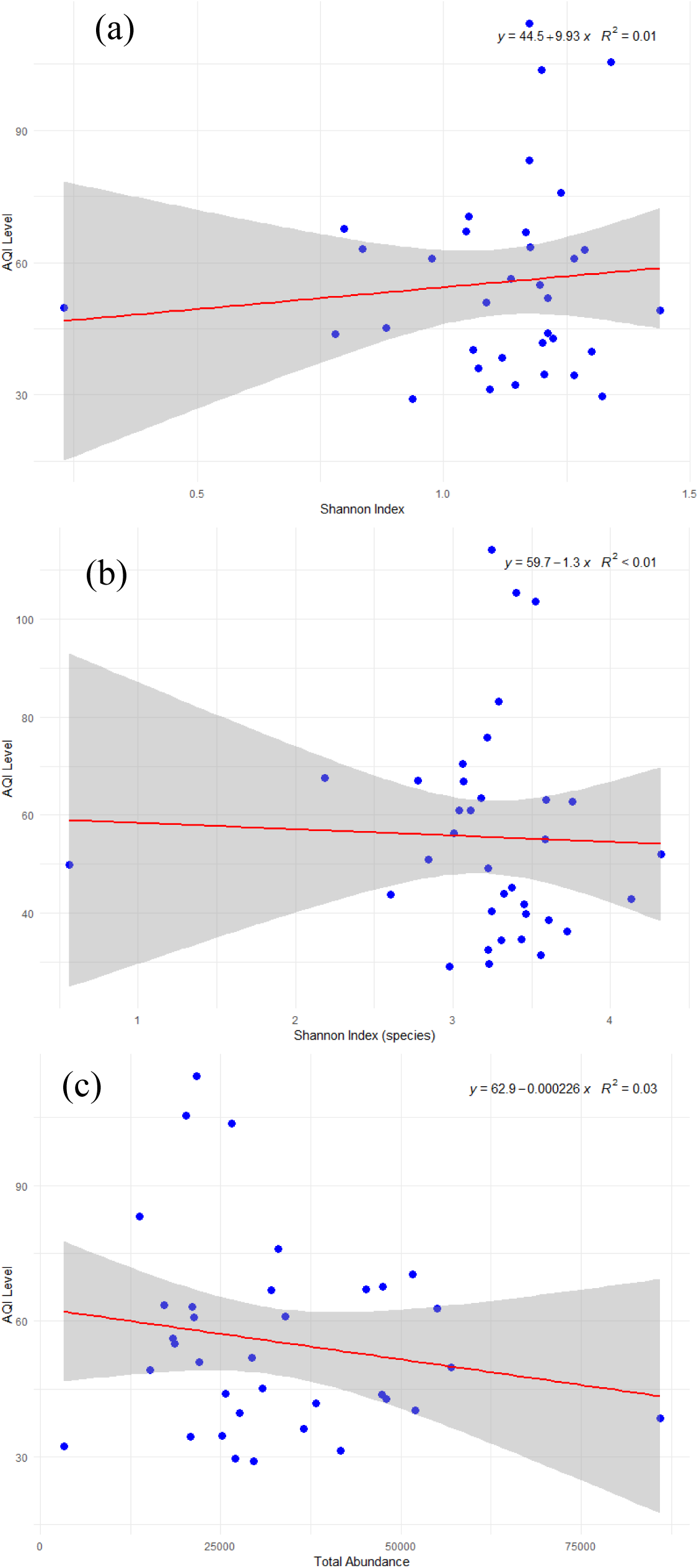
Linear regression of AQI level and (a) phylum-level Shannon index, (b) ASV-level Shannon index, and (c) total bacterial abundance

One possible explanation for this discrepancy could be the difference in how AQI is defined across countries. In our study, the AQI level is determined by concentrations of PM_2.5_, PM_10_, CO, O_3_, SO_2_, NO, and NO_X_, whereas the AQI regulation by Chilean government is only based on PM_2.5_ concentration. The definitional differences might contribute to the divergent results. Furthermore, since Taipei City generally has better air quality compared to Temuco, where Acuña et al. conducted their research. This difference might have introduced a range restriction effect that obscures potential relationships between air quality and bacterial diversity [35].

In contrast, the AQI range in Wang et al. study is comparable to ours, allowing us to rule out range restriction as the cause of the inconsistency between our results and theirs. Notably, our study included a larger sample size, which might lend greater confidence to our findings. Taken together, these results suggest that AQI levels may have minimal or negligible correlation with air bacterial diversity in environments with relatively good air quality.

Additionally, we found minor correlation between AQI level and total bacterial abundance (R^2^ = 0.03), as shown in Fig. 4c. This finding differs from former study that reported positive correlations during specific air pollution events such as Asian dust events and cyclical haze events [36, 37]. Again, the limited variability in AQI levels within our study area may explain this discrepancy.

### 3.3 Spatial distribution of airborne SARS-CoV-2 aerosols

Table 2 shows the detection results of SARS-CoV-2 in each sampling site. In this study, we define negative pressure isolation room as “hot zone”, other sampling sites in hospital as “warm zone”, and the sampling sites in campus area as “cold zone”. This definition is based on the probability that one could expose to the viruses in that place. As Table 2 indicates, the detection rate is highest in hot zone (87.5%), followed by warm zone (58.3%) and cold zone (25.0%).

The detection rate at the nurse station near negative pressure isolation room was higher when there were COVID-19 patients in the wards compared to when no patients were admitted, which were 100% and 33.3%, respectively. This suggests that the presence of patients might contribute to environmental biohazards. Although several nosocomial infection control measures are in place, such as (i) mandatory use of personal protective equipment (PPE) for staff entering isolation rooms, (ii) a strict policy prohibiting PPE from being removed from the ward, and (iii) a buffer isolation area with double doors between the isolation ward and the nursing station, our findings indicate that environmental biocontamination may still occur in adjacent non-isolation areas [38–40]. Additionally, there was one positive sample in nurse station while no COVID-19 patients admission. This might imply a potential infection of healthcare workers or a complete and exhaustive disinfection measurement was undone.

For hospital lobby, positive samples were observed in daytime and nighttime, while the former was more than the latter. This might be due to the high human traffic during daytime. Despite the fact that the sample collection period was near the end of pandemic and the authorities had mask-wearing policy when entering medical facilities, our study suggested that medical institutions remain high-risk area for viral exposure compared to other settings.

Fig. S4 shows the C_t_ value of samples in each zone. The viral load was highest in hot zone, followed by warm zone and cold zone. This imply that hospital setting not only has higher chance to expose to SARS-CoV-2 virus but also has higher concentration of viral load in the air.

### 3.4 Factors influencing bacterial community and viral distribution

We used RDA to investigate the association between bacterial structure and environmental factors. The considered environmental variables included PM_1_ chemical species (i.e., ion, metal, OC/EC), meteorological factors (e.g., wind speed, temperature, relative humidity, etc.) and gaseous pollutants (i.e., CH_4_, THC, CO, O_3_, NO, NO_x_). The RDA biplot is shown in Fig. 5. In the figure, the blue arrows indicate bacterial phyla and the black arrows indicate environmental factors included in the model. The greater absolute value of the cosine of the angle between the arrows suggest a stronger correlation between them. This means that the arrow in the same and opposite direction indicate a positive and negative correlation, respectively, while an angle close to right angle imply a weak correlation [27, 41].

**Fig. 5.**
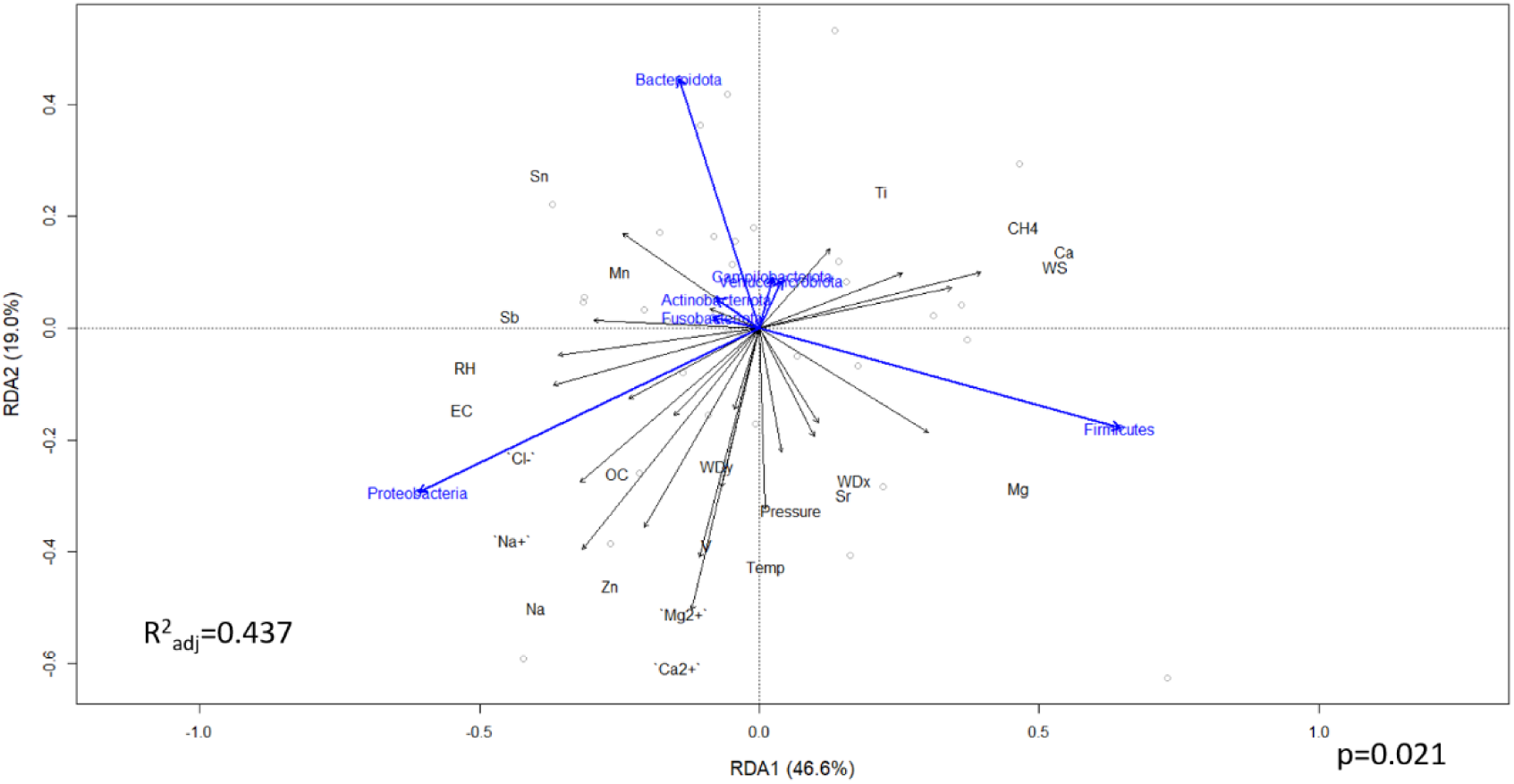
RDA biplot illustrating the relationships between environmental variables and bacterial phyla. Blue arrows represent bacterial phyla, and black arrows represent environmental variables, including PM₁ chemical species, meteorological parameters, and gaseous pollutants. Environmental variables with variance inflation factors (VIF) > 20 were excluded. WS, wind speed; WDx/WDy, cosine and sine components of wind direction; Temp, temperature; RH, relative humidity

The adjusted R^2^ is 0.437, suggesting that 43.7% of the bacterial community variance could be explained by the environmental variables selected in the model. From the biplot, *Bacteroidota* has positive correlation with Sn and Mn, and has negative correlation with Sr, barometric pressure and temperature. Although some of the findings have not been reported by other studies, a similar study about soil bacterial community also supported the positive correlation between Mn and *Bacteroidota* [42].

*Firmicutes* has positive correlation with Mg, Ca, wind speed and CH_4_, while has negative correlation with Sn, Mn, Sb and relative humidity. Since Mg and Ca are crustal elements and contained in road dusts, this might imply that *Firmicutes* have correlate with local and primary pollutants [43, 44]. On the other hand, *Proteobacteria* has positive correlation with OC, EC, Na^+^, Cl^-^ and relative humidity and negative with Ti, Ca, wind speed and CH_4_. From past experiences, EC primarily comes from combustion activities [45, 46], and Na^+^ and Cl^-^ mainly represent sea salt sources [47, 48]. However, the sampling site is far from any incinerators and sea, hence this result might suggest that *Proteobacteria* have higher association with transported pollutants.

We further conducted variance partitioning analysis (VPA), as shown in Fig. 6. This analysis could help better understand the explained variance of each environmental variables [49]. The results indicated that, when controlling meteorological factors and gaseous pollutants, PM_1_ chemical species itself could explain 40.67% of bacterial community variance. Similarly, meteorological factors can explain 43.14% of variance of bacterial community, while gaseous pollutants explained only 9.58%. Furthermore, the three categories mutually explained 14.16% of variance. Notably, when we excluded gaseous pollutants from the all-factor RDA model, the adjusted R^2^ decreased to 0.335. While the individual explanatory power of gaseous pollutants appeared lower compared to PM_1_ and meteorological variables, this reduction in explanatory value implies that their influence on bacterial community structure may still be meaningful and should not be overlooked.

**Fig. 6.**
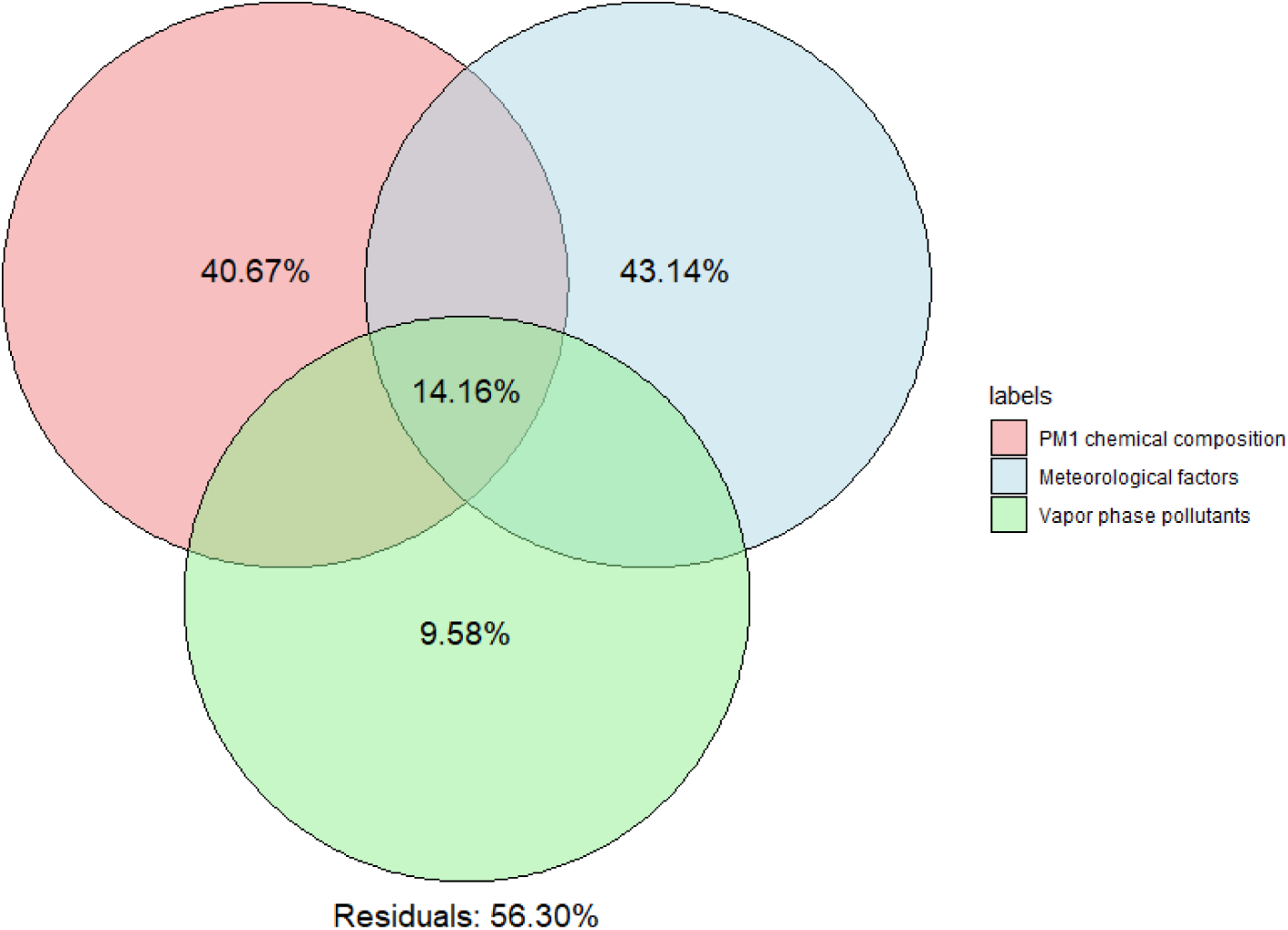
Variance partitioning analysis (VPA) based on redundancy analysis (RDA) showing the relative contributions of PM₁ chemical composition, meteorological factors, and gaseous pollutants to the variation in bacterial community composition.

Additionally, when we add extra environmental variables, oxidative potential (OP), to all-factor RDA, we observed that OP_V_ primarily showed positive correlation with *Proteobacteria*, while OP_M_ was positively correlated with *Firmicutes* and negatively correlated with *Proteobacteria*. However, the adjusted R^2^ further declined to 0.294. One possible explanation for this decrease is that including OP may have led to the removal of other environmental variables due to high collinearity, some of which may have had stronger explanatory power. It is also possible that the relationship between OP and bacterial community is not linear, limiting the ability of RDA, a linear model, to fully capture these associations. Furthermore, the VPA results showed that OP itself could only explain 1.19% of bacterial community variance, suggesting that OP had a relatively minor impact on bacterial community structure (Fig. S5).

We further conducted RDA including only PM_1_ chemical species, meteorological factors and gaseous pollutants, respectively, as shown in Fig. S6. In terms of PM_1_ chemical species, comparing with the variance partitioning in Fig. 6, the explanatory power in PM_1_-only RDA decreased to 12.0%. This implied that meteorological factors and gaseous pollutants might modulate or enhance the impact of PM_1_ chemical species on bacterial community structure. A similar pattern was observed for meteorological factors and gaseous pollutants when analyzed individually. These findings collectively implied that PM_1_ chemical species, meteorological factors, and gaseous pollutants might interact in a synergistic manner to shape airborne bacterial community.

At genus level, we also conducted RDA and mainly focused on pathogenic bacterial genera, including *Bacteroides* (phylum *Bacteroidota*), *Helicobacter* (phylum *Campylobacterota*), *Corynebacterium* (phylum *Actinobacteria*), *Streptococcus* (phylum *Firmicutes*), and *Escherichia/Shigella*, *Pseudomonas*, *Proteus*, *Klebsiella*, *Acinetobacter* (phylum *Proteobacteria*) (Fig. 7). The results showed that PM_1_ chemical composition, meteorological factors, and gaseous pollutants could explain 52.2% of variance of pathogenic bacterial community. *Escherichia*/*Shigella* and *Bacteroides* could be best explained by the RDA axes. From the RDA biplot, *Escherichia*/*Shigella* has positive correlation with Na, Zn, and barometric pressure, while has negative correlation mainly with northerly winds. *Bacteroides* has positive correlation with Sn, EC, Mn, Cl^-^, and Ti, and has negative correlation with wind speed. In addition, it shows minimal correlation with wind direction. These results might imply that *Bacteroides* thrive better under conditions with lower air movement, potentially due to reduced dispersion or environmental disturbance.

**Fig. 7.**
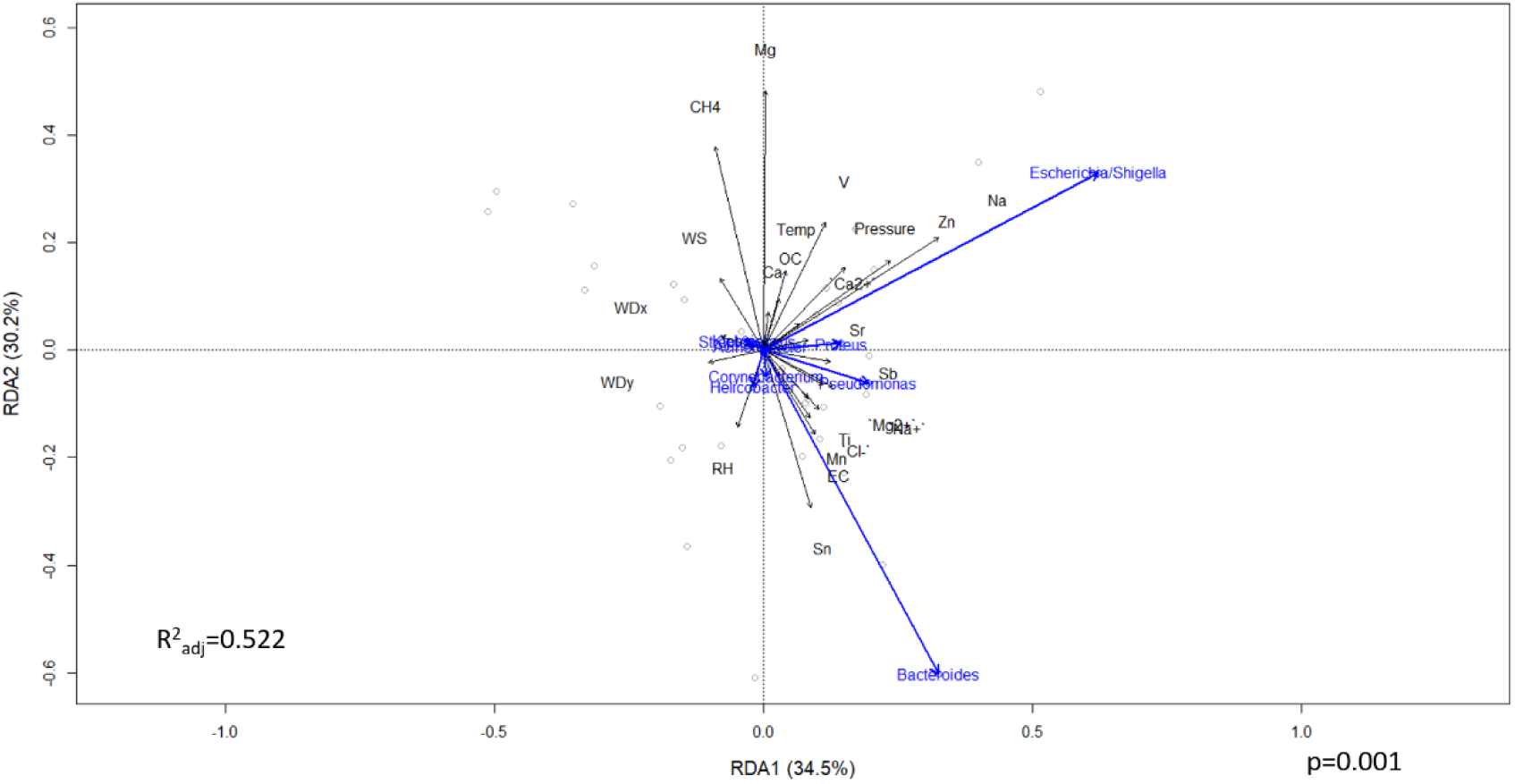
RDA biplot illustrating the relationships between environmental variables (PM₁ chemical composition, meteorological parameters, and gaseous pollutants) and pathogenic bacterial genera. WS, wind speed; WDx/WDy, cosine and sine components of wind direction; Temp, temperature; RH, relative humidity

As for VPA results, the outcome is analogous with phylum-level RDA. PM_1_ chemical species solely could explain 47.48% of bacterial variance, while meteorological factors and gaseous pollutants explain 43.25% and 8.04%, respectively (Fig. 8a). Again, when adding OP into the RDA model, the adjusted R^2^ decreased from 0.552 to 0.324, and VPA result showed that the explained bacterial variance of OP is -0.91% (Fig. 8b). This negative value could be interpreted as an indication of “no explanatory power” beyond random chance [50]. This result suggested a similar implication that OP might have minor impact on bacterial community structure.

**Fig. 8.**
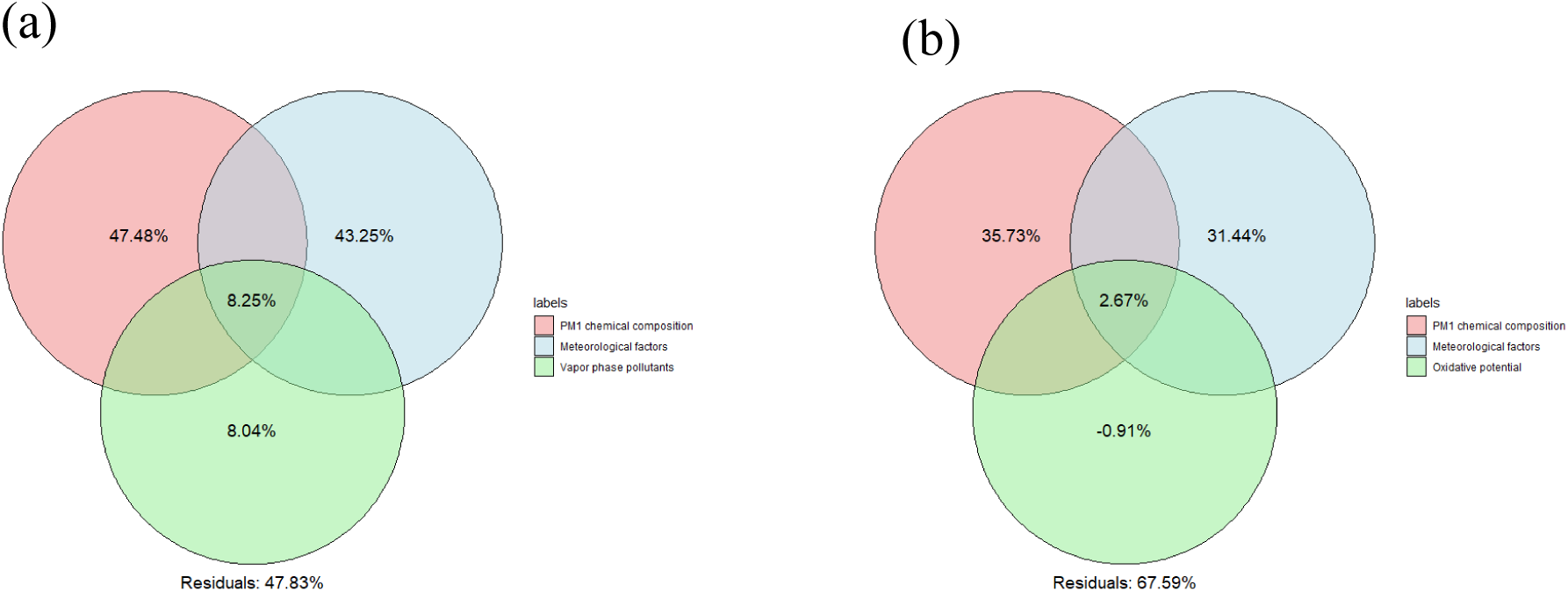
Variance partitioning analysis (VPA) based on redundancy analysis (RDA) illustrating the relative contributions of environmental variable groups to the variation in pathogenic bacterial community composition. The RDA models included (a) PM₁ chemical species, meteorological factors, and gaseous pollutants, and (b) the same variables with the addition of oxidative potential (OP)

Furthermore, from correlation analyses (Fig. S7a), we found that vanadium (V) and antimony (Sb) have the highest positive correlation with total bacterial abundance. One possible explanation of the positive correlation between Sb and bacterial abundance could be the co-emission effect from traffic-related sources. Sb is primarily associated with anthropogenic activities, such as automotive brake abrasion, fly ash from waste incinerator, and traffic emission [51, 52]. Simultaneously, high-traffic environments significantly contribute to the resuspension of road dust, which has been shown to increase airborne bacterial abundance [53]. Considering that our sampling site is near a major arterial road in Taipei City, the co-occurrence could result in a positive correlation between Sb and bacterial abundance.

For vanadium, it is widely recognized as a tracer for heavy fuel oil combustion, particularly from marine vessels [54, 55]. In parallel, bacterial abundance was also found to be positively associated with easterly winds (Fig. S7c). Considering that Port of Keelung, one of Taiwan’s largest international seaports, is located to the northeast of Taipei City, it is plausible that air masses arriving from this direction carry ship-emitted V-rich aerosols toward the urban sampling site [56]. In addition, Taipei municipal wastes incinerator and Second funeral parlor (crematory) are located to the near east of the sampling site. These could probably generate Sb-rich aerosols and also be carried to the sampling site.

Bacterial abundance was positively correlated with OP_V_ and negatively correlated with OP_M_ (Fig. S7b), with Spearman’s ρ = 0.29 and -0.31, respectively. The positive association with OP_V_ may be partially explained by the fact that higher OP_V_ are linked to greater PM load (Spearman’s ρ = 0.35, figure not shown), which could provide more niches for bacterial colonization. In contrast, higher OP_M_ implies stronger toxicity, which could impose environmental stress on airborne bacteria, potentially leading to a reduction in their abundance.

Lastly, we found temperature also appeared to be relatively positively correlated with bacterial abundance. This trend might reflect the general tendency of higher temperatures to promote bacterial growth, thereby contributing to increased abundance [57].

Having examined the environmental determinants of bacterial composition in the air, the next step is to assess how other factors influence the detection rate of airborne viruses. We sought to assess the influence of human traffic on viral detection, and thus we estimated human traffic flow in both warm and cold zones. Human traffic was defined as high when peak flow exceeded 100 individuals per hour and as low when the flow was below this threshold. Stepwise logistic regression was then employed to identify factors associated with viral detection, with candidate variables including temperature, relative humidity, wind speed, barometric pressure, and peak human traffic. The final model retained only peak human traffic as a significant predictor (*p*=0.007), suggesting that peak human traffic was significantly associated with viral detection. To validate this association, we conducted a contingency table analysis. Given the limited sample size, Fisher’s exact test was applied to evaluate the relationship between peak human traffic and viral detection. The analysis revealed a statistically significant correlation (*p*=0.038), further supporting the role of human traffic as a contributing factor to environmental viral presence. This result implied that crowd gathering places are potential viral hotspots.

We may suppose that this is because the higher volume of people increases the likelihood that some individuals carry and shed the virus, thus elevating the chance of environmental detection. If individuals who carry the virus move between locations, they may unintentionally transport the virus to new areas, promoting its spatial spread. Therefore, human mobility does not merely affect the intensity of viral presence in a single place, it also plays a critical role in widening the transmission border across different regions [58, 59].

## Supporting information

graphical abstract

## Data Availability

All data referred to in the manuscript are available from the corresponding author upon reasonable request. Publicly available air quality and meteorological data used in this study can be accessed from the Taiwan Ministry of Environment open data platform.

## Acknowlegements

This research was supported by the National Science and Technology Council, Taiwan (NSTC 113-2111-M-A49-001). The authors gratefully acknowledge the contributions of all team members involved in field sampling, chemical analysis, and epidemiological data processing. We also appreciate the constructive feedback provided by the editor and anonymous reviewers, which significantly strengthened the final version of this manuscript.

## CRedit authorship contribution statement

Yu Hsiang Chen: Conceptualization, Methodology, Formal analysis, Data curation, Writing – original draft. Po Jui Chen: Conceptualization, Methodology, Formal analysis, Data curation, Writing – original draft. Kun Ta Chou: Data curation, Validation. Hsiang Ling Ho: Investigation, Resources. Kai Yu Hsu: Data curation, Visualization, Software. Kung Hung Chieh: Data curation, Investigation. Ta Chih Hsiao: Investigation, Resources. Mey Jy Jeng: Resources. Tse Min Wei: Investigation, Resources. Hsiao Chi Chuang: Writing – review & editing. Kai Hsien Chi: Supervision, Project administration, Funding acquisition, Writing – review & editing.

## Data availability

All data used in this study were obtained from publicly available sources. Details of the data sources and data processing procedures are described in the Methods section.

