## Supplementary Materials for "Chemical and Biological Characteristics of PM₁-Associated Aerosols and Airborne Viruses in Hospital and Campus Environments during the Post-COVID Period"

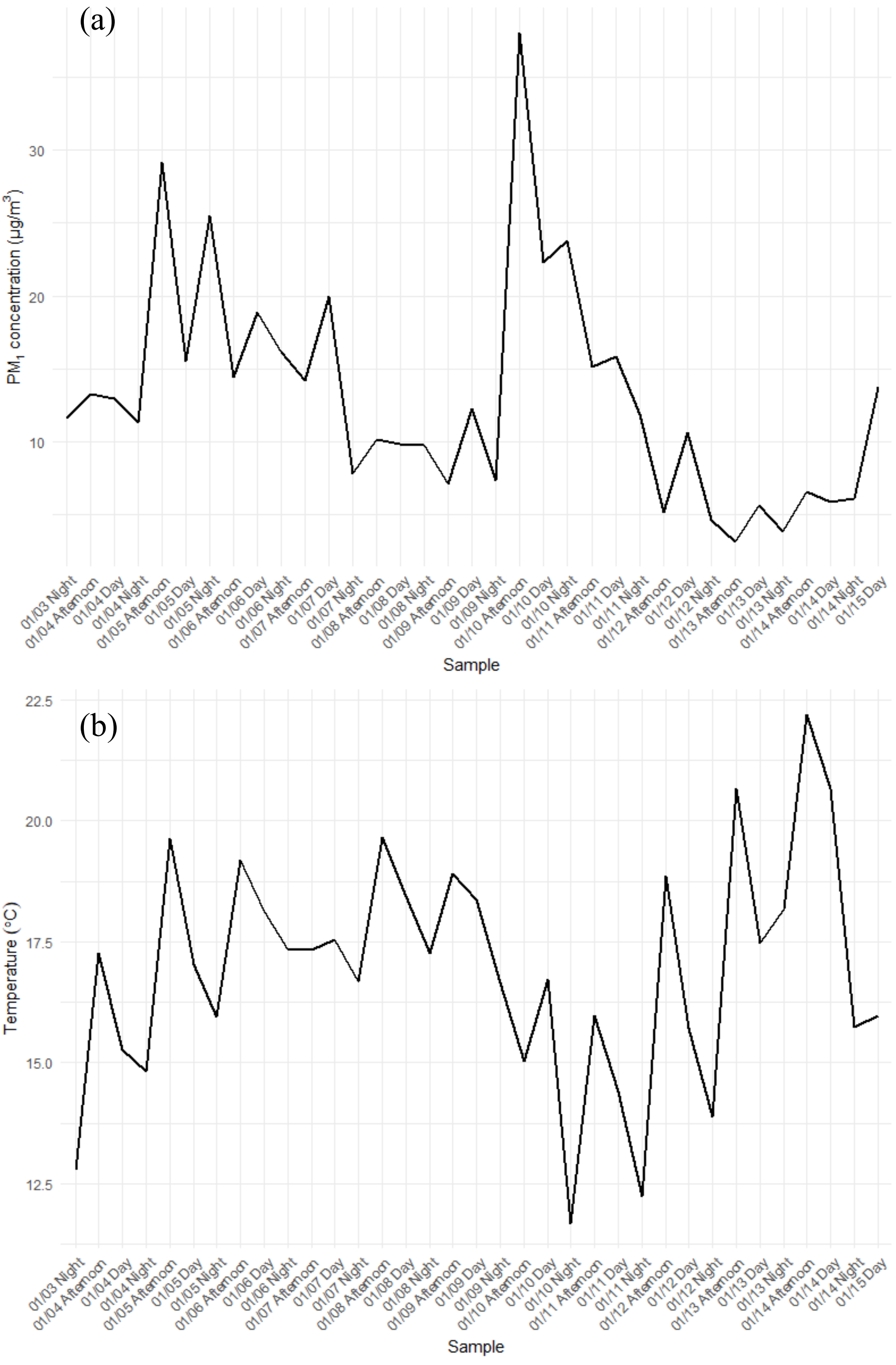


Fig. S1. Temporal variations of (a) $PM_1$ concentrations and (b) ambient temperature

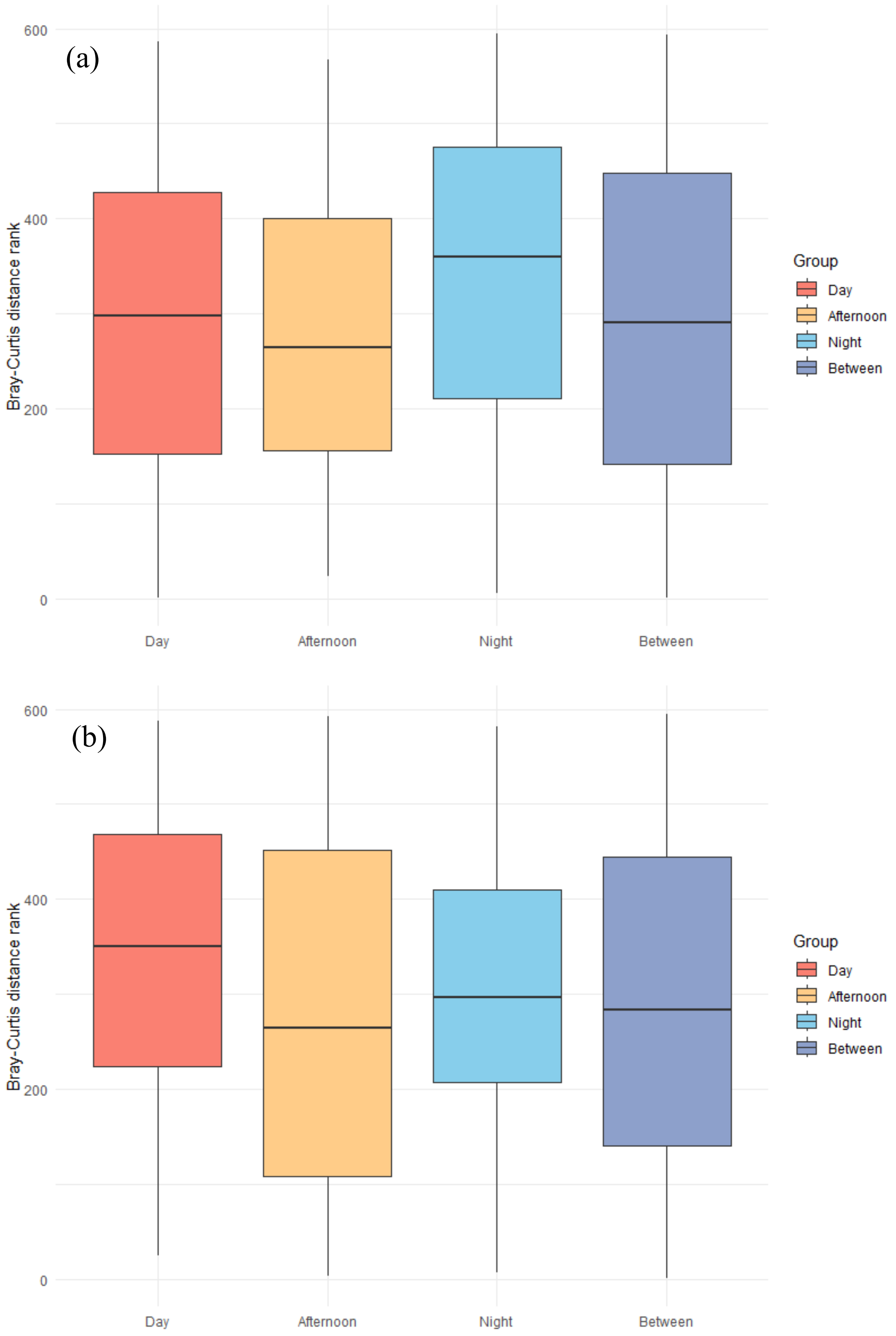


during the sampling period
Fig. S2 Analysis of similarity (ANOSIM) of (a) ionic species and (b) metal distribution. No significant differences were observed among day, afternoon, and night samples

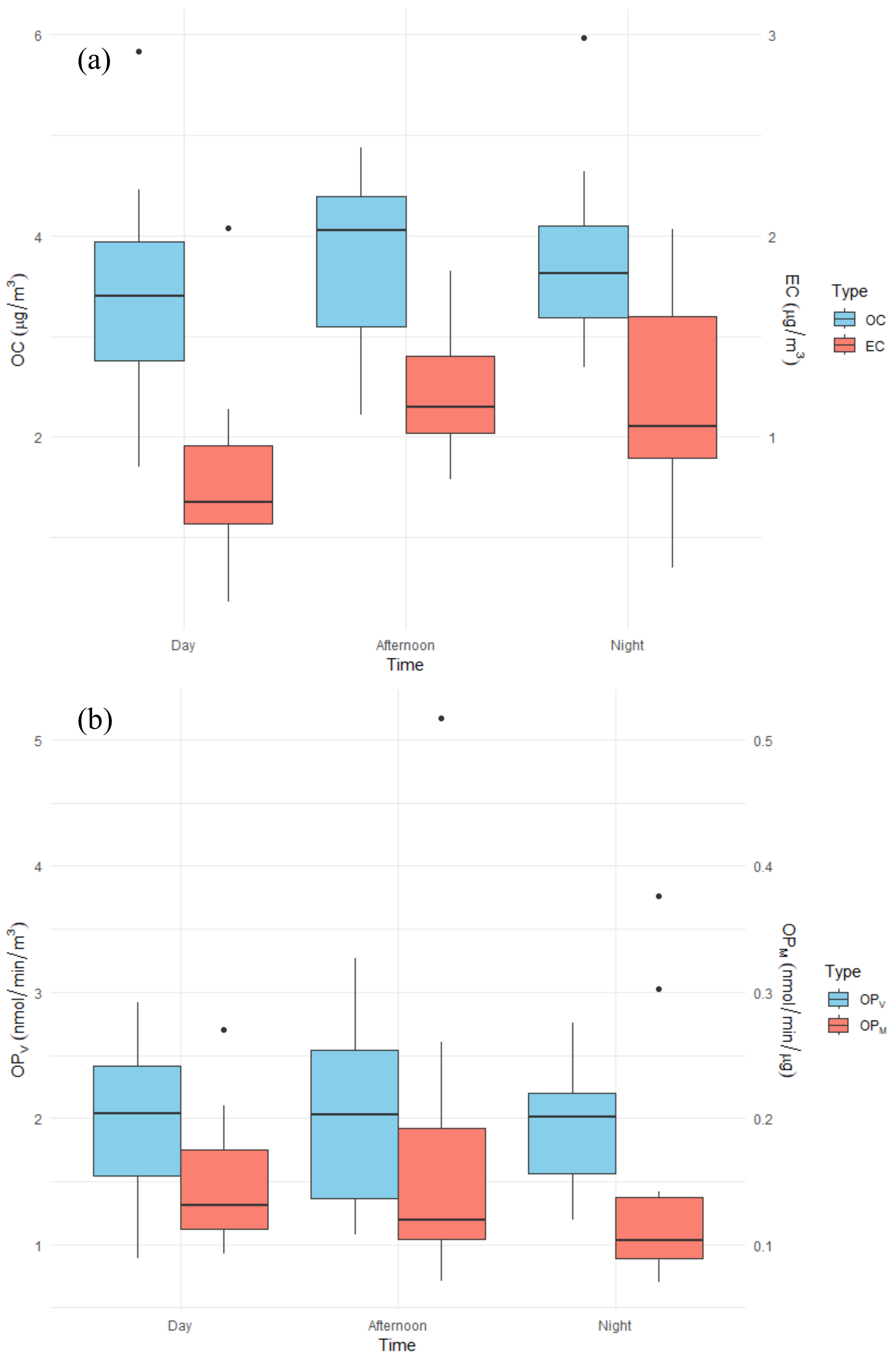


Fig. S3 Diurnal variations of (a) OC/EC and (b) OPV/OPM in day, afternoon, and nighttime samples

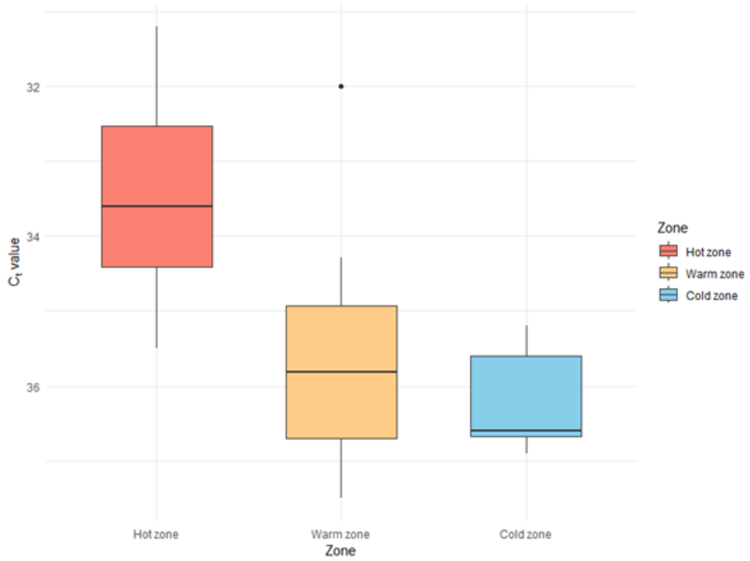


Fig. S4. Ct values of samples across different zones

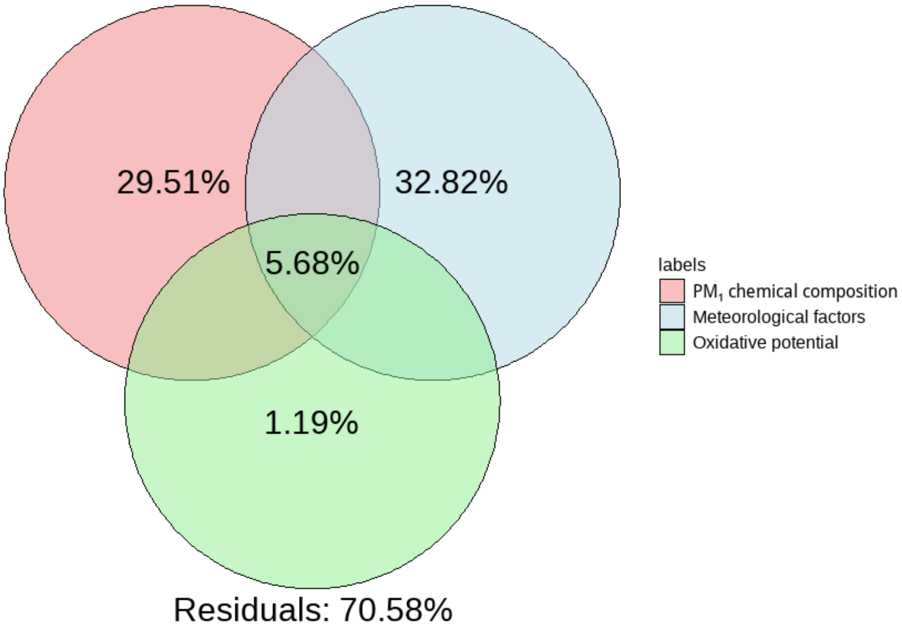


Fig. S5. Variance partitioning analysis (VPA) based on redundancy analysis (RDA) including oxidative potential (OP). All vapor-phase pollutant variables were excluded due to high variance inflation factors (VIFs) after the inclusion of OP variables

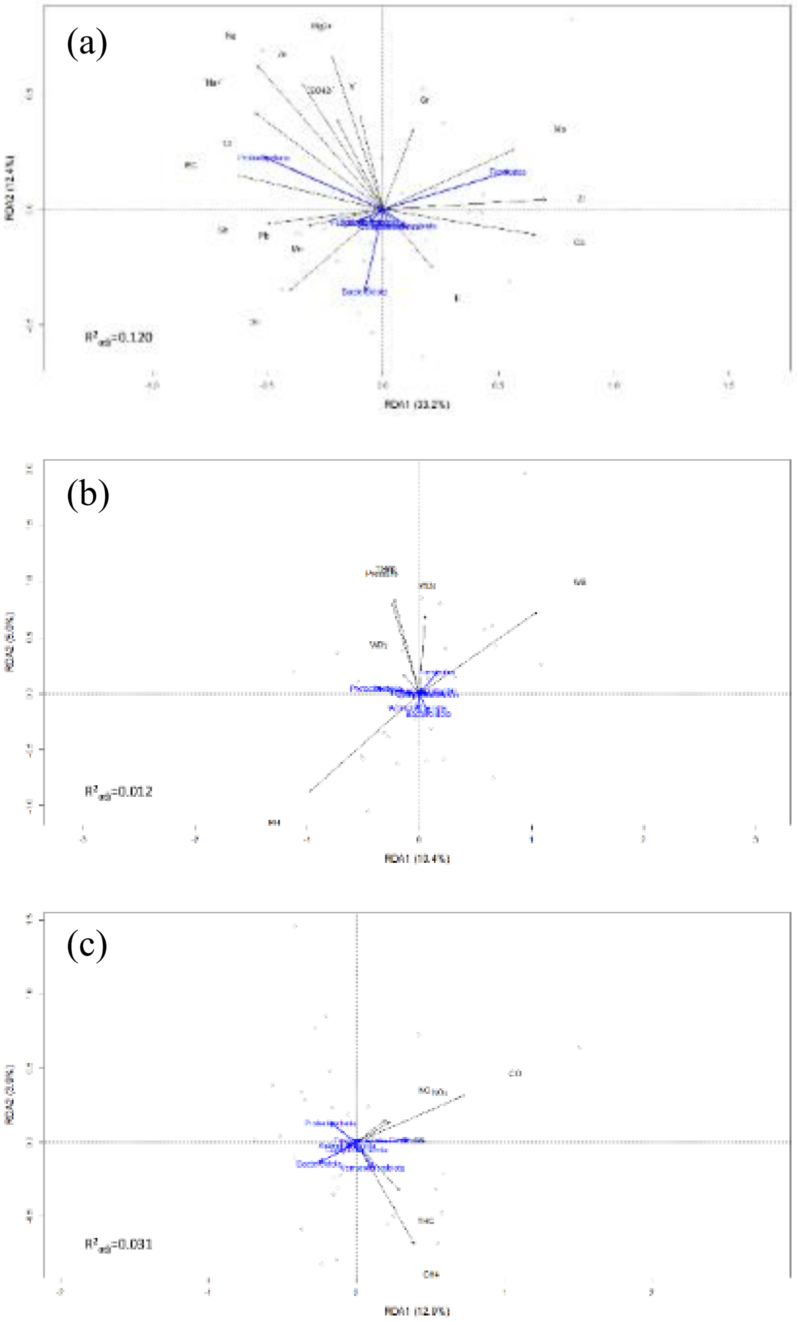


Fig. S6. RDA biplot illustrating the relationships between bacterial community composition and (a) $PM_1$ chemical species, (b) meteorological factors, and (c) gaseous pollutants

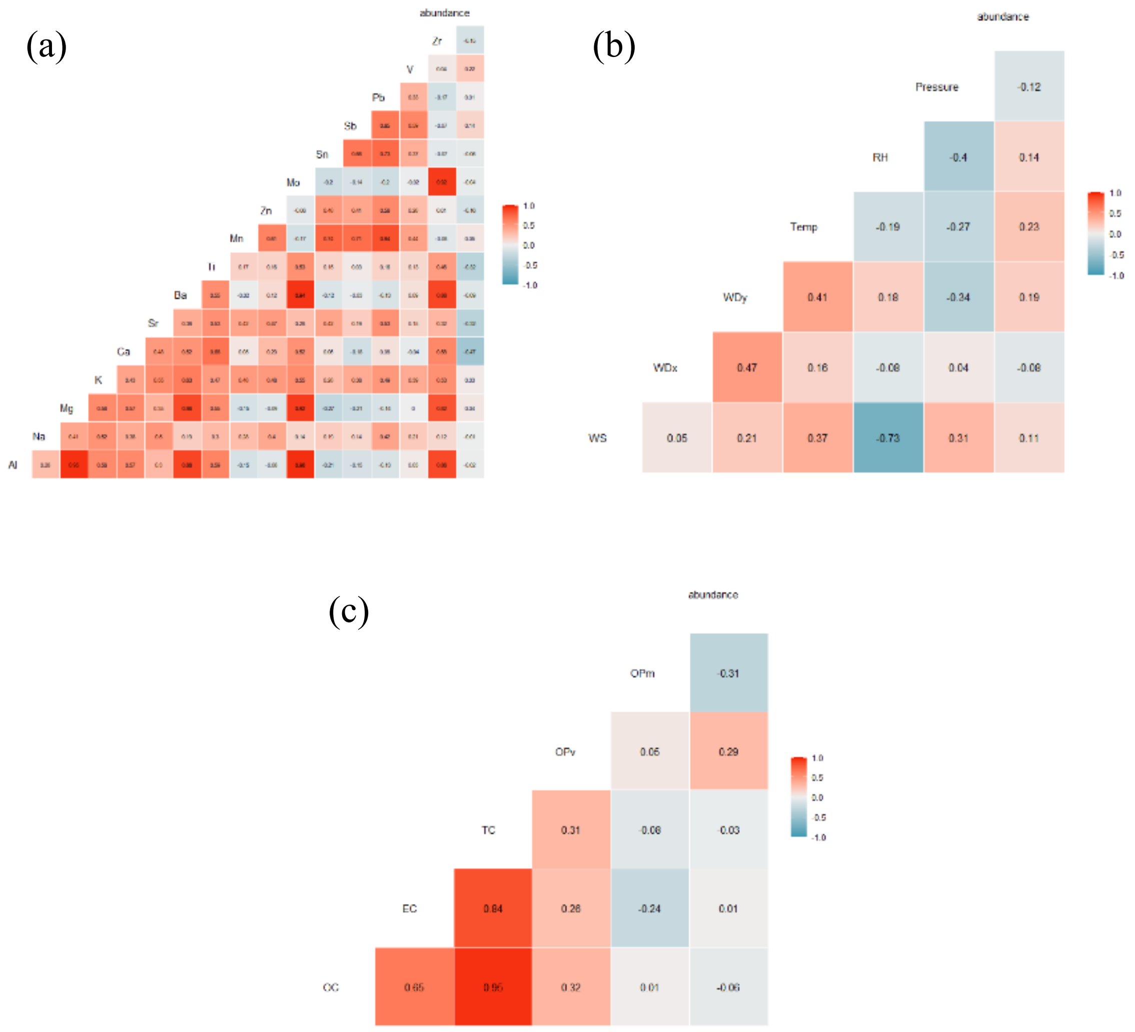


Fig. S7. Spearman correlation of total bacterial abundance and (a) PM metal concentration, (b) OC/EC and $OP_V/OP_M$, (c) meteorological factors. V and Sb have the highest positive correlation with bacterial abundance, as well as northerly winds and temperature. RH: relative humidity; Temp: temperature; WDx/WDy: cosine/sine of wind direction; WS: wind speed
