## Supplementary material for "Chemical and Biological Characteristics of PM₁-Associated Aerosols and Airborne Viruses in Hospital and Campus Environments during the Post-COVID Period": graphical abstract

**Aerosol $PM_1$**
Temporal dynamics of $PM_1$-associated aerosols in campus and hospital environments

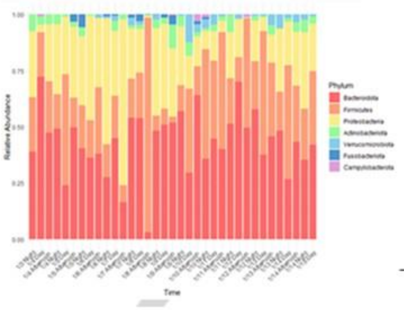


**Bioaerosol Composition**
Environmental drivers shaping bioaerosol community composition

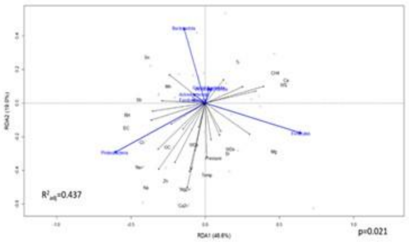


**Environmental Viral Detection**
Coverage-based assessment of airborne viral signals in campus and hospital settings

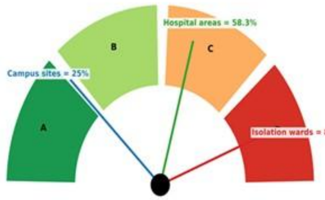


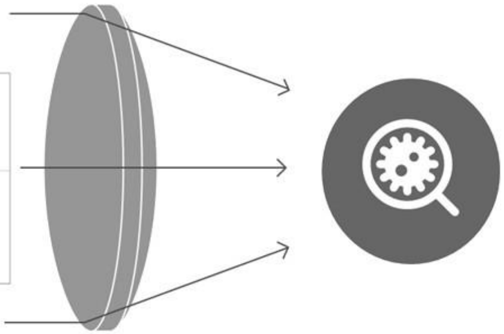

**Cross-Setting Surveillance of $PM_1$-Associated Bioaerosols and Environmental Viral Detection**
